# Sequential rTMS-tACS Neuromodulation Is Associated with Increased Resting-State Theta Activity in Fibromyalgia: A Randomized Controlled Pilot Study

**DOI:** 10.64898/2026.10.27.26364954

**Authors:** Öznur Buran, Behzad Rostami Sani, Daniel Cabello, Pedro Montoya

## Abstract

**Background:** Sequential application of repetitive transcranial magnetic stimulation (rTMS) and transcranial alternating current stimulation (tACS) may induce neurophysiological changes within sensorimotor and pain-related networks. However, the resting-state EEG effects of this combined approach in fibromyalgia remain unknown.

**Methods:** In this exploratory randomized controlled pilot study, 10 adults with fibromyalgia were assigned to active stimulation (n = 5) or a no-stimulation control group (n = 5). Active participants received one or two sessions of 10-Hz rTMS over the left primary motor cortex (1,000 pulses/session), followed by 10 daily sessions of bilateral 10-Hz tACS (1 mA, 20 min). Six-minute eyes-closed resting-state EEG recordings were obtained before and after the intervention or an equivalent control interval. Power spectral density was quantified in the delta, theta, alpha, and beta bands and analyzed after natural-log transformation using mixed repeated-measures ANOVAs across frontal, central-centroparietal, and posterior regions of interest (ROIs), followed by separate regional Time × Group analyses.

**Results:** Theta power increased significantly in the active group but not in controls, yielding a significant Time × Group interaction (F(1,8) = 15.09, p = .005, ηp² = .654). Follow-up analyses showed increased theta power in frontal (p = .015), central-centroparietal (p = .006), and posterior (p = .012) regions. However, the Time × ROI × Group interaction was not significant (p = .059), providing no evidence of regional differences in group-dependent changes. A significant Time × Group interaction was also observed for delta power (p = .019), but not for alpha or beta power.

**Conclusions:** Sequential motor-cortex rTMS followed by repeated bilateral 10-Hz tACS was associated with increased resting-state theta activity across frontal, central-centroparietal, and posterior scalp regions in fibromyalgia. Given the small sample size, post hoc ROI selection, lack of sham stimulation, and absence of component-specific control conditions, these findings should be considered hypothesis-generating and require replication in adequately powered sham-controlled studies.

## Introduction

Fibromyalgia is a chronic pain condition characterized by widespread pain, fatigue, sleep disturbances, cognitive complaints, and affective symptoms (Clauw, 2014). Although its pathophysiology remains incompletely understood, converging evidence points to altered central processing of nociceptive and non-nociceptive information and mechanisms consistent with central sensitization (López-Solà et al., 2017; Sluka & Clauw, 2016; Woolf, 2011). Neuroimaging studies further indicate abnormalities within pain-related and sensorimotor networks, including disrupted connectivity within the pain network (Cifre et al., 2012) and reduced communication between sensorimotor, insular, and thalamocortical regions (Flodin et al., 2014). Together, these findings suggest that altered sensorimotor processing contributes to pain hypersensitivity in fibromyalgia.

Consistent with this view, resting-state EEG studies have identified abnormalities in oscillatory activity, functional connectivity, and network organization in fibromyalgia and other chronic pain conditions (González-Roldán et al., 2016; González-Villar et al., 2020; Diniz et al., 2024; Zebhauser et al., 2023). However, no resting-state EEG measure has demonstrated sufficient consistency or specificity to serve as a biomarker of chronic pain (Ta Dinh et al., 2019; Zebhauser et al., 2023; Zortea et al., 2021). Although a recent systematic review reported an overall tendency toward reduced delta, theta, and alpha activity and increased beta activity in fibromyalgia (Silva-Passadouro et al., 2024), findings remain heterogeneous, including reports of increased slow-frequency activity in fibromyalgia and other chronic pain conditions (González-Roldán et al., 2016; Lopes et al., 2024). This variability likely reflects differences in methodology, participant characteristics, and clinical status and suggests that neuromodulatory effects should be examined across multiple frequency bands and scalp regions rather than assuming a uniform EEG signature (Silva-Passadouro et al., 2024; Thut et al., 2017).

Repetitive transcranial magnetic stimulation (rTMS) of the primary motor cortex (M1) can modulate cortical excitability and distributed pain-related networks. In fibromyalgia, clinical improvement following M1-rTMS has been associated with changes in thalamic, insular, cingulate, frontal, and parietal connectivity, together with reduced pain and normalization of motor-cortical excitability measures (Argaman et al., 2022a, 2022b; Molero-Chamizo et al., 2023). Transcranial alternating current stimulation (tACS), in contrast, applies weak sinusoidal currents that interact with ongoing neural oscillations in a frequency-dependent manner (Herrmann et al., 2013; Vogeti et al., 2022). Stimulation at 10 Hz is of particular interest because it overlaps with the sensorimotor mu rhythm, although evidence regarding repeated bilateral 10-Hz sensorimotor stimulation remains limited, particularly in fibromyalgia (Bernardi et al., 2021; Lafleur et al., 2021).

The present protocol combined repeated tACS with preceding M1-rTMS. Because both interventions interact with the physiological state of the targeted network, their sequential application may produce effects that differ from those of either modality alone (Neuling et al., 2013; Ridding & Ziemann, 2010; Hosseinian et al., 2021). However, evidence for such interactions derives mainly from concurrent or phase-synchronized paradigms in healthy individuals, and their relevance to sequential clinical interventions remains uncertain. Moreover, neuromodulation studies have reported post-stimulation changes outside the frequency range of stimulation, justifying an exploratory examination of multiple EEG frequencies and scalp regions.

Therefore, this exploratory randomized controlled pilot study investigated resting-state EEG changes associated with a sequential neuromodulation protocol consisting of high-frequency M1-rTMS followed by repeated bilateral 10-Hz sensorimotor tACS in individuals with fibromyalgia. Given the limited and heterogeneous literature, analyses were conducted across multiple frequency bands and scalp regions without assuming *a priori* that stimulation effects would be restricted to alpha activity or to electrodes directly overlying the stimulation sites.

## Methods

### Participants

Ten adults with fibromyalgia participated in the study (nine women and one man; mean age = 50.5 years, SD = 9.1). Participants were recruited through advertisements at a local hospital and in collaboration with the *Associació Mallorquina de Fibromiàlgia, Síndrome de Fatiga Crònica/EM, Sensibilitat Química Múltiple i Electrohipersensibilitat* (AMASC). All participants underwent screening for contraindications to non-invasive brain stimulation and EEG recording, and none met any exclusion criteria. The study was approved by the Ethics Committee of the Regional Government of the Balearic Islands (reference GOIBE549363/2026), and all participants provided written informed consent before taking part.

As this was an exploratory pilot study, no formal *a priori* power calculation was conducted. The sample size was determined pragmatically based on the number of eligible participants who could be recruited and complete the protocol during the predefined study period. The study was designed to provide preliminary estimates of intervention-related neurophysiological effects and to inform the design and sample size planning of future adequately powered trials. Recruitment and data collection concluded at the end of the predefined study period. Participants’ sociodemographic characteristics and questionnaire scores are presented in Table 1.

**Table 1.**
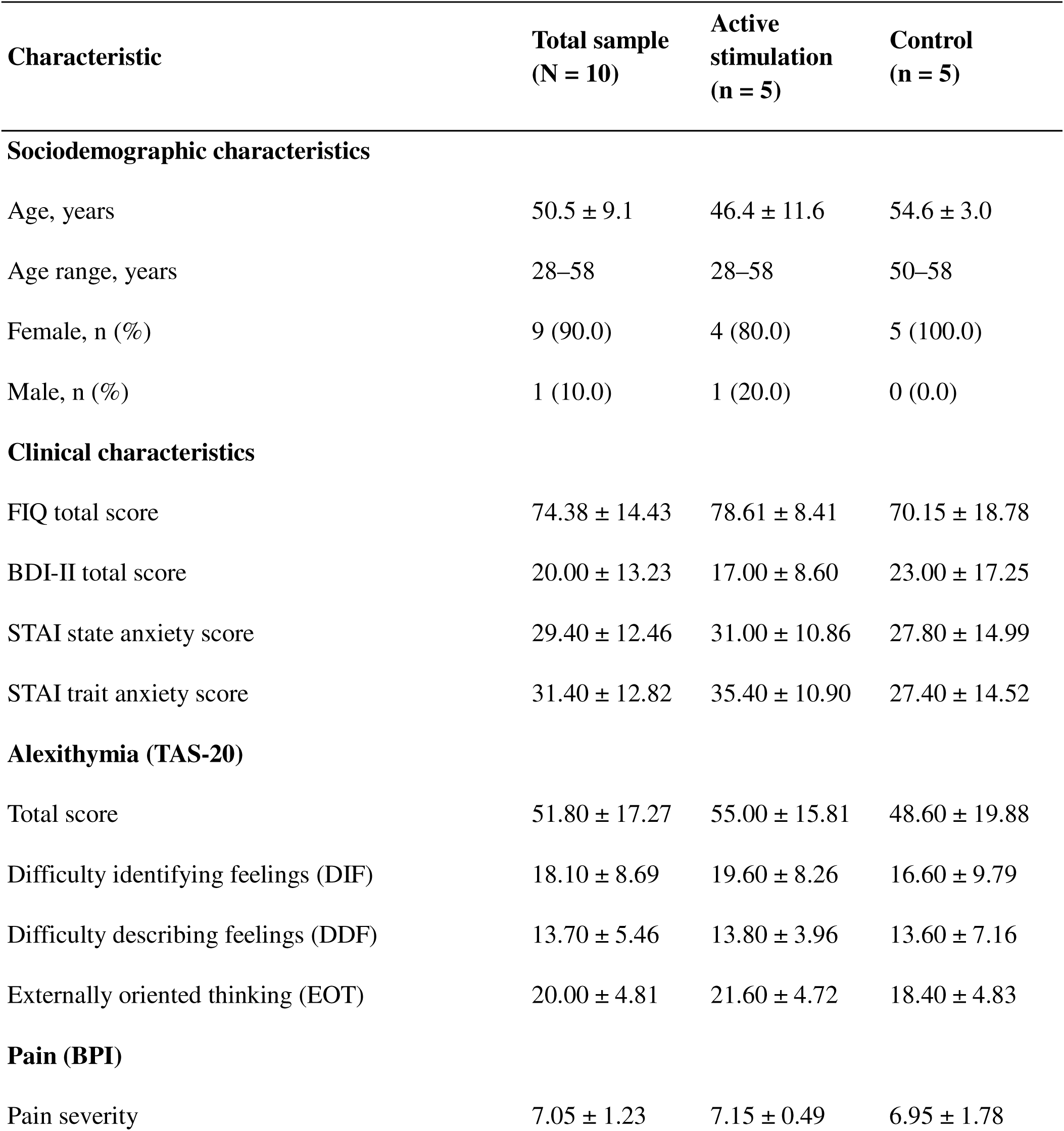
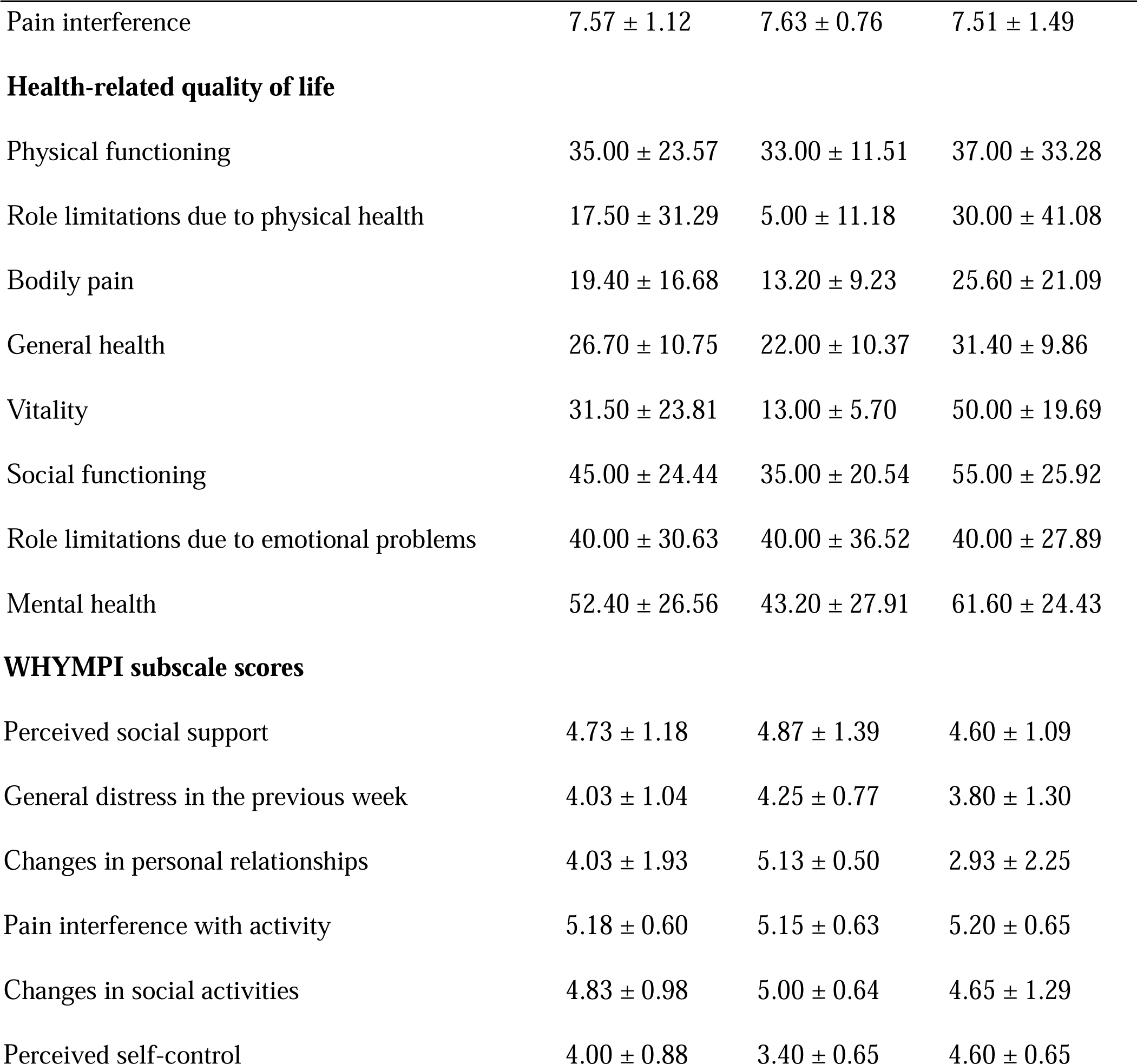
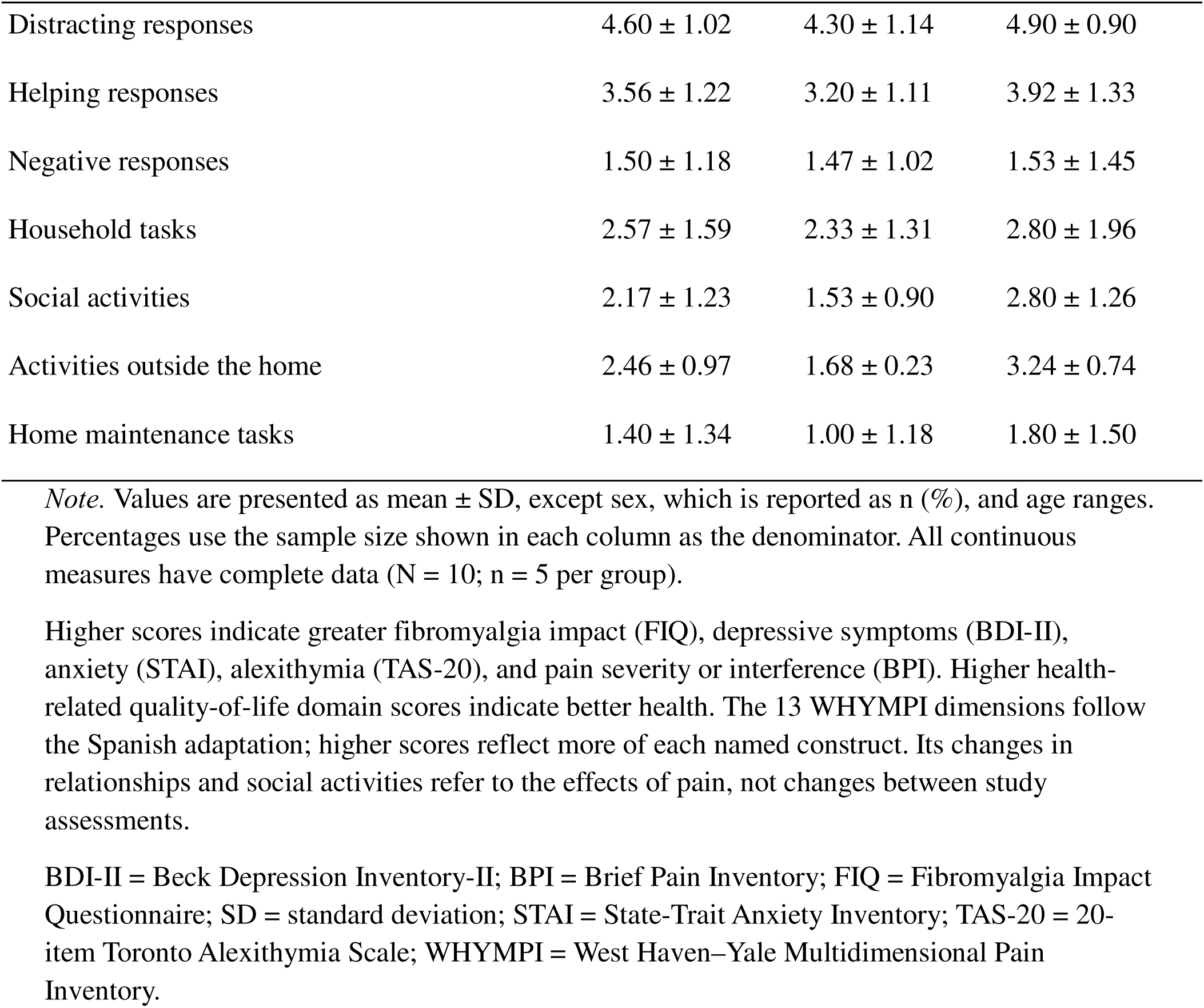
Sociodemographic and clinical characteristics of participants with fibromyalgia.

### Materials and Instruments

Prior to the experimental session, participants completed a structured clinical interview and a battery of self-report questionnaires administered online using Google Forms (Google LLC, Mountain View, CA, USA). All instruments were administered in their validated Spanish versions. The clinical interview collected information on sociodemographic characteristics, medical history, and fibromyalgia-related clinical features. The impact of fibromyalgia on health status and daily functioning was assessed with the Fibromyalgia Impact Questionnaire (FIQ; Burckhardt et al., 1991; Rivera & González, 2004). The FIQ evaluates physical functioning, work-related difficulties, and core fibromyalgia symptoms, including pain, fatigue, stiffness, sleep disturbance, anxiety, depression, and overall well-being. Pain severity and pain-related interference with daily activities were assessed using the Brief Pain Inventory (BPI; Cleeland & Ryan, 1994; Badia et al., 2003). Health-related quality of life was measured with the 36-Item Short Form Health Survey (SF-36; Ware & Sherbourne, 1992; Alonso et al., 1995), which assesses eight domains of physical and mental health. Alexithymia was assessed using the 20-Item Toronto Alexithymia Scale (TAS-20; Bagby et al., 1994; Martínez-Sánchez, 1996), which evaluates difficulty identifying feelings, difficulty describing feelings, and externally oriented thinking. The multidimensional experience of chronic pain was assessed with the West Haven– Yale Multidimensional Pain Inventory (WHYMPI; Kerns et al., 1985; Ferrer-Pérez et al., 1993), which measures pain-related interference, perceived social responses to pain, and participation in everyday activities. Depressive symptom severity was assessed using the Beck Depression Inventory-II (BDI-II; Beck et al., 1996; Sanz et al., 2003), whereas anxiety was assessed using the State–Trait Anxiety Inventory (STAI; Spielberger et al., 1983; Spielberger et al., 1982), which evaluates both situational anxiety (state) and the relatively stable tendency to experience anxiety (trait). Questionnaire scores are summarized in Table 1.

### Experimental Design and Procedure

This exploratory pilot study employed a randomized, parallel-group controlled design with two resting-state EEG assessments: baseline (t0) and post-intervention or follow-up (t1) (Figure 1). Participants were assigned by simple randomization to either an active-stimulation group (n = 5) or a non-stimulation control group (n = 5). Because the control condition did not include sham stimulation, neither participants nor the study assessor were blinded to group allocation.

**Figure 1.**
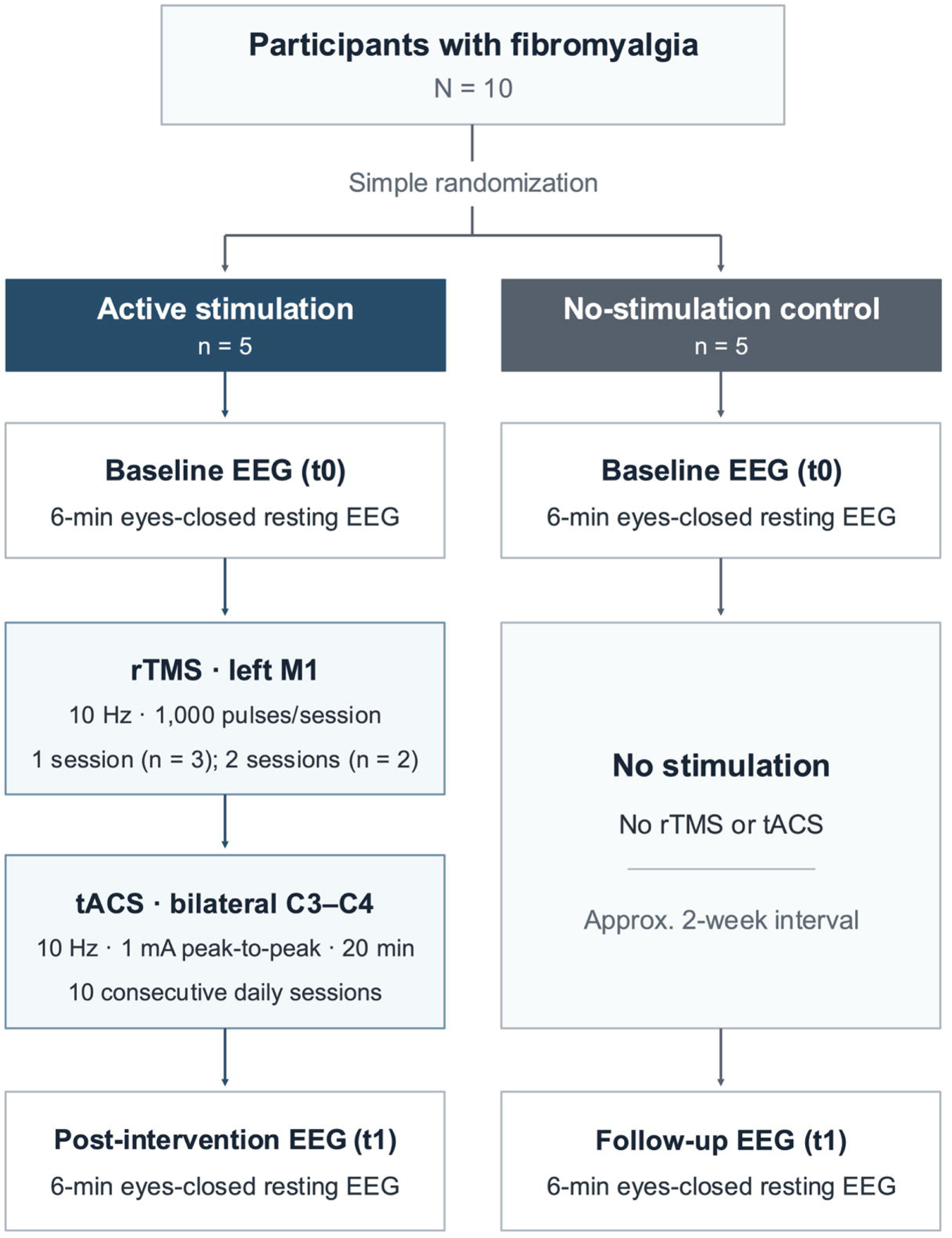
Experimental design of the randomized controlled pilot study. Ten participants with fibromyalgia were randomly assigned to an active-stimulation group (n = 5) or a no-stimulation control group (n = 5). Both groups underwent a 6-min eyes-closed resting-state electroencephalography (EEG) recording at baseline (t0). The active group received one or two sessions of 10-Hz repetitive transcranial magnetic stimulation (rTMS) over the left primary motor cortex (M1; 1,000 pulses/session), followed by 10 consecutive daily sessions of 10-Hz transcranial alternating current stimulation (tACS; 1 mA peak-to-peak, 20 min) delivered over C3 and C4. A second resting-state EEG recording was obtained after completion of the intervention (t1). The control group received no stimulation and underwent a second EEG recording approximately two weeks after baseline.

At baseline, all participants underwent a 6-min eyes-closed resting-state EEG recording (t0). Participants assigned to the active-stimulation group subsequently received a sequential neuromodulation protocol consisting of rTMS followed by repeated tACS. Owing to feasibility constraints, rTMS exposure was not identical across participants: two received two rTMS sessions administered on consecutive days, whereas three received a single session. Following the final rTMS session, all participants completed ten bilateral tACS sessions delivered on ten consecutive days. A second 6-min eyes-closed resting-state EEG recording (t1) was obtained after completion of the final tACS session. Participants assigned to the control group received neither rTMS nor tACS during the study period. Instead, they returned approximately two weeks after the baseline assessment to undergo a second 6-min eyes-closed resting-state EEG recording (t1), providing a time-matched comparison for the active-stimulation group.

### EEG Acquisition and Preprocessing

Resting-state EEG was recorded using an actiCHamp Plus amplifier and a 64-channel actiCAP slim system (Brain Products GmbH, Germany). Sixty-one scalp electrodes were positioned according to the international 10-10 system. Signals were acquired at 1000 Hz with CPz as the online reference and FPz as ground. Vertical electrooculographic (EOG) activity was recorded from electrodes placed above and below the left eye. Electrode impedances were maintained below 10 kΩ. Participants completed a 6-min eyes-closed resting-state recording while seated comfortably and instructed to remain awake.

EEG data were processed offline in BrainVision Analyzer 2. Following González-Roldán et al. (2025), the first 60 s were discarded and the remaining 300 s segmented into consecutive 1-s epochs. Data were cleaned using a 50-Hz notch filter, filtered (0.05-35 Hz; Butterworth) and corrected using the Gratton-Coles ocular-artifact correction procedure (Gratton et al., 1983). Epochs containing amplitudes exceeding ±100 µV, voltage differences greater than 100 µV, or voltage steps exceeding 100 µV were rejected automatically. Participants with fewer than 75% artifact-free epochs were excluded.

Power spectral density (PSD) was estimated using a fast Fourier transform with a 10% Hanning window and zero-padding to 1024 points (frequency resolution = 0.98 Hz). PSD values were averaged across retained epochs and computed for delta (1-4 Hz), theta (4-8 Hz), alpha (8-12 Hz), and beta (12-30 Hz) frequency bands.

Regional analyses were conducted using three exploratory scalp regions of interest (ROIs) defined after visual inspection of the topographical distribution of intervention-related changes. The frontal ROI included Fp1, Fp2, AF3, AF4, AF7, AF8, AFz, F1, F2, F5, F6, F7, F8, and Fz (14 electrodes). The central-centroparietal ROI included FC1, FC2, FC3, FC4, FC5, FC6, FCz, C1, C2, C3, C4, C5, C6, Cz, CP1, CP2, CP3, CP4, CP5, and CP6 (20 electrodes). The posterior ROI included P1, P2, P3, P4, P5, P6, P7, P8, Pz, PO3, PO4, PO7, PO8, POz, O1, O2, and Oz (17 electrodes). The arithmetic mean PSD was calculated across electrodes within each ROI, and the natural logarithm of the resulting mean was used for statistical analysis. Because ROI selection was data-driven, regional analyses should be considered exploratory and interpreted cautiously (Kriegeskorte et al., 2009).

### rTMS Stimulation Protocol

High-frequency rTMS was delivered over the left primary motor cortex (M1), a target previously investigated for pain modulation in fibromyalgia (Passard et al., 2007; Lee et al., 2012; Argaman et al., 2022a, 2022b). The approximate M1 location was identified using standard 10-20 EEG-system landmarks, and the stimulation hotspot was defined as the scalp position where single-pulse stimulation elicited the most consistent visible muscle twitch in the contralateral right hand.

Resting motor threshold was determined by visual observation as the lowest stimulator intensity producing a visible hand-muscle response in at least 5 of 10 consecutive trials (Badran et al., 2019). Stimulation intensity was set at 100% of resting motor threshold. Each session consisted of 20 trains delivered at 10 Hz (50 pulses per train; 1,000 pulses/session). Trains lasted 5 s and were separated by 8-s intertrain intervals, resulting in a total stimulation time of approximately 4 min.

Stimulation was delivered using an Apollo TMS system and an air-cooled figure-of-eight coil (MAG & More GmbH, Munich, Germany). Owing to feasibility constraints, rTMS exposure varied slightly among participants: three received a single session and two received two sessions on consecutive days. Following completion of the rTMS phase, all participants proceeded to the tACS intervention.

### tACS protocol

tACS was delivered using a battery-powered constant-current stimulator (DC-STIMULATOR PLUS, neuroConn GmbH, Ilmenau, Germany). Two circular electrodes (20 mm diameter; ∼3 cm² contact area) were positioned over C3 and C4 according to the international 10-10 EEG system, approximately corresponding to the left and right sensorimotor cortices. Conductive paste was used to ensure stable contact, and electrode impedance was maintained below 5 kΩ throughout stimulation.

A 10-Hz sinusoidal current (1 mA peak-to-peak) was applied for 20 min, including 30-s ramp-up and ramp-down periods. Participants completed 10 sessions delivered on consecutive days following the rTMS phase. During stimulation, participants were seated comfortably and instructed to rest quietly. Stimulation parameters were based on previous studies employing bilateral sensorimotor tACS (Lafleur et al., 2021; Wach et al., 2013) and were consistent with established safety recommendations for low-intensity transcranial electrical stimulation (Antal et al., 2017; Woods et al., 2016).

### Statistical analyses

Statistical analyses were conducted using IBM SPSS Statistics 29 (IBM Corp., Armonk, NY, USA). Separate mixed-design repeated-measures ANOVAs were performed for log-transformed delta, theta, alpha, and beta power, with Time (t0, t1) and ROI (frontal, central-centroparietal, posterior) as within-subject factors and Group (active stimulation, control) as a between-subject factor. The models included all main effects, two-way interactions, and the Time × ROI × Group interaction. Additional exploratory mixed-design ANOVAs were conducted separately for each ROI, with Time as the within-subject factor and Group as the between-subject factor, to examine regional longitudinal changes. Bonferroni-adjusted pairwise comparisons were used to examine differences between ROIs, changes across time within each group, and group differences at each time point. Statistical significance was set at p < .05, and effect sizes were expressed as partial eta squared (ηp²).

## Results

### Delta (1–4 Hz)

The mixed ANOVA showed significant main effects of Time, F(1,8) = 6.14, p = .038, ηp² = .434, and ROI, F(2,16) = 59.66, p < .001, ηp² = .882, as well as a significant Time × Group interaction, F(1,8) = 8.58, p = .019, ηp² = .518. Bonferroni-corrected comparisons indicated higher delta power in the frontal ROI than in the central-centroparietal and posterior ROIs (both p < .001), with no significant difference between the latter two regions (p = 1.000). Delta power decreased significantly from baseline (t0) to follow-up (t1) in the control group (p = .005), whereas no significant change was observed in the active-stimulation group (p = .757). The Time × ROI × Group interaction was not significant, F(1.19, 9.54) = 2.07, p = .183, ηp² = .206 (Greenhouse–Geisser corrected). Separate exploratory ANOVAs showed significant Time × Group interactions in the frontal (p = .027), central-centroparietal (p = .038), and posterior (p = .035) ROIs. Follow-up comparisons indicated significant decreases in the control group across all three regions (p = .002, .016, and .017, respectively), with no significant changes in the active-stimulation group.

### Theta (4–8 Hz)

Theta-band power showed a significant Time × Group interaction, F(1,8) = 15.09, p = .005, ηp² = .654, whereas the Time × ROI × Group interaction was not significant, F(2,16) = 3.39, p = .059, ηp² = .298. A significant main effect of ROI was also observed, F(2,16) = 57.99, p < .001, ηp² = .879. Bonferroni-corrected comparisons indicated higher theta power in the frontal ROI than in the central-centroparietal (p < .001) and posterior (p = .001) ROIs, with higher power also observed in the posterior than in the central-centroparietal ROI (p = .032).

Follow-up analyses indicated a significant increase in theta power from baseline (t0) to post-intervention (t1) in the active-stimulation group when averaged across the three ROIs (p = .005), whereas no significant change was observed in the control group (p = .129). Separate exploratory ANOVAs showed significant Time × Group interactions in the frontal, F(1,8) = 9.62, p = .015, ηp² = .546, central-centroparietal, F(1,8) = 13.90, p = .006, ηp² = .635, and posterior ROIs, F(1,8) = 11.43, p = .010, ηp² = .588. Bonferroni-adjusted comparisons indicated significant increases in theta power in the active-stimulation group across the frontal (p = .015), central-centroparietal (p = .006), and posterior (p = .012) regions, whereas no significant changes were observed in any ROI in the control group. Figure 2 presents the resting-state EEG power spectra across the frontal, central-centroparietal, and posterior regions at baseline (t0) and post-intervention or follow-up (t1) for the active-stimulation (Figure 2A) and control (Figure 2B) groups, respectively.

**Figure 2.**
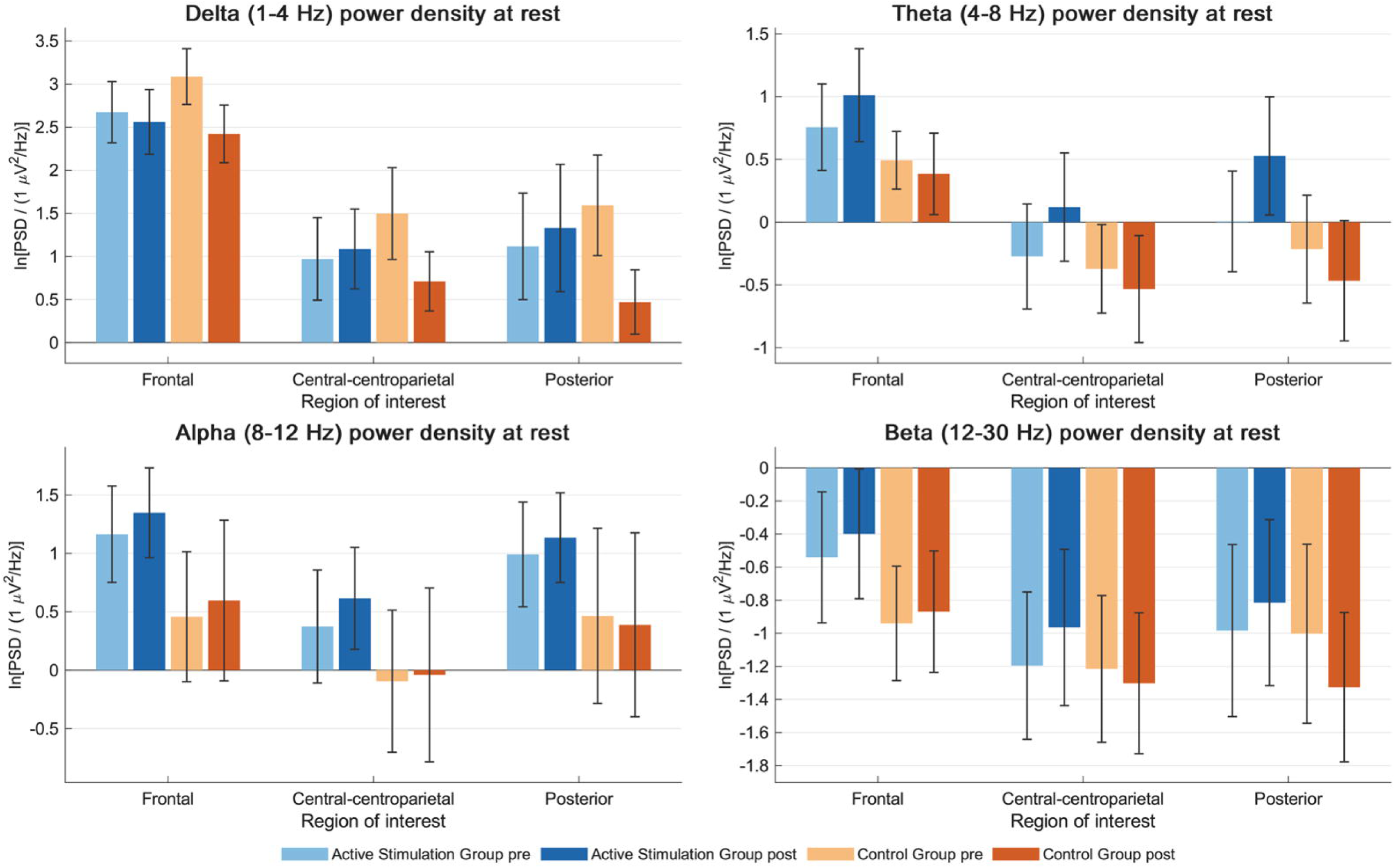
Resting-state EEG power spectra across frontal, central–centroparietal, and posterior regions before and after intervention. Grand-average power spectral density (PSD; µV²/Hz) across the 4–30 Hz frequency range in the active stimulation group (A; n = 5) and the control group (B; n = 5). Black and red lines denote pre-intervention (t□) and post-intervention or follow-up (t□), respectively. For each participant, spectra were averaged across electrodes within each region of interest and then averaged across participants within each group.

### Alpha(8–12 Hz)

A significant main effect of ROI was observed, F(2,16) = 23.17, p < .001, ηp² = .743. Bonferroni-corrected comparisons indicated higher alpha power in the posterior (p = .002) and frontal (p < .001) ROIs than in the central-centroparietal ROI, with no significant difference between the frontal and posterior regions (p = .936). No significant main effects of Time, F(1,8) = 1.06, p = .333, ηp² = .117, or Group, F(1,8) = 0.65, p = .443, ηp² = .075, were found. All two-way interactions and the Time × ROI × Group interaction were nonsignificant (all p ≥ .332). Separate exploratory ANOVAs also showed no significant Time × Group interactions in the frontal (p = .876), central-centroparietal (p = .455), or posterior (p = .350) ROIs. Bonferroni-adjusted comparisons showed no significant pre/post changes within either group in any region.

### Beta (12–30 Hz)

A significant main effect of ROI was observed, F(2,16) = 16.62, p < .001, ηp² = .675. Bonferroni-corrected comparisons indicated higher beta power in the frontal ROI than in the central-centroparietal (p < .001) and posterior (p = .045) ROIs, with no significant difference between the latter two regions (p = .316). No significant main effects of Time, F(1,8) = 0.17, p = .689, ηp² = .021, or Group, F(1,8) = 0.23, p = .646, ηp² = .028, were observed. Likewise, none of the two-way interactions or the Time × ROI × Group interaction reached significance (all p ≥ .109). Separate exploratory ANOVAs showed no significant Time × Group interactions in the frontal (p = .597), central-centroparietal (p = .098), or posterior (p = .112) ROIs. Bonferroni-adjusted comparisons showed no significant pre/post changes within either group in any region. Figure 3 illustrates the mean log-transformed power spectral density across the three ROIs for all four frequency bands at baseline (t0) and post-intervention or follow-up (t1) in the active stimulation and control groups.

**Figure 3.**
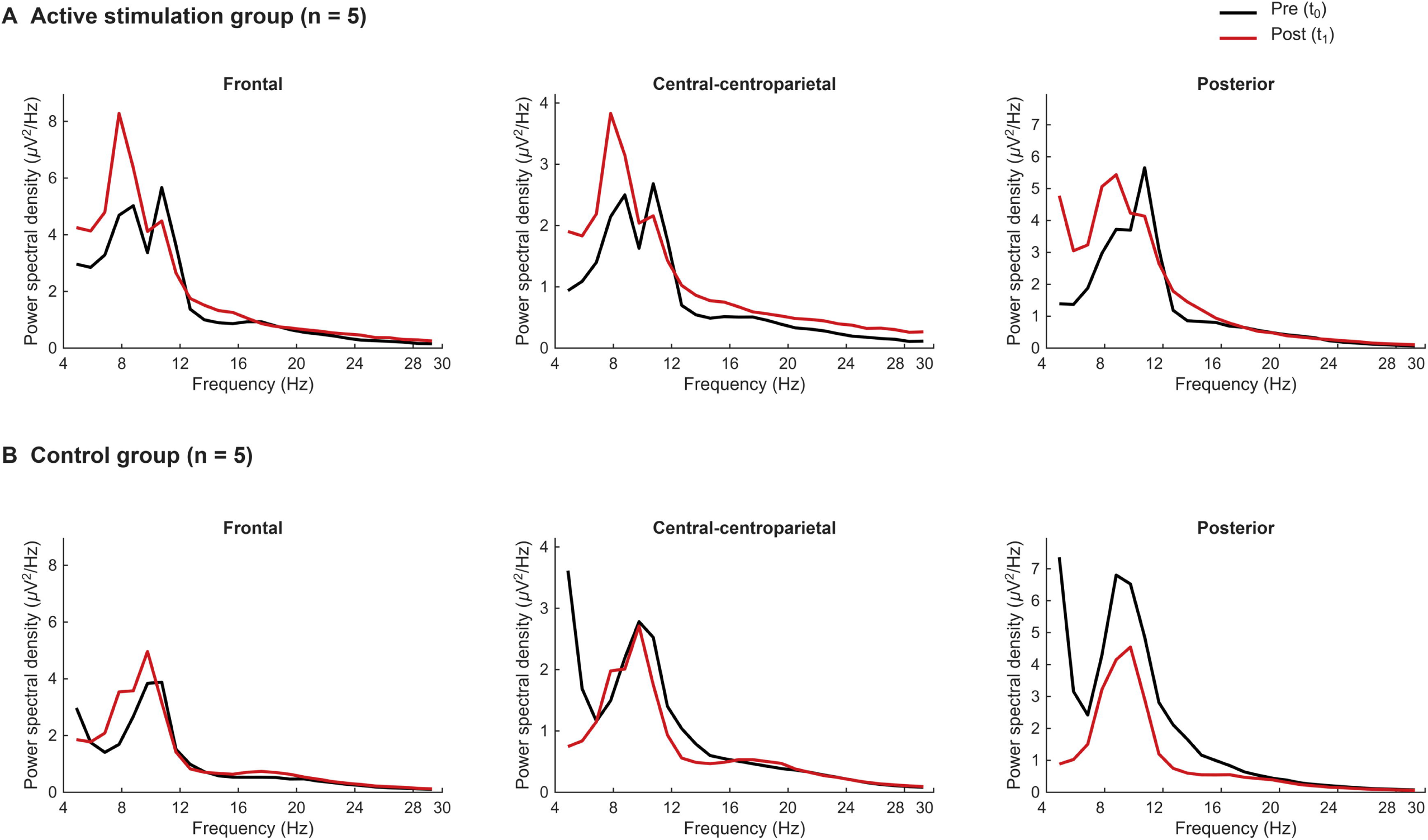
Regional changes in resting-state EEG power spectral density following intervention. Mean natural-log-transformed power spectral density (PSD) across delta (1–4 Hz), theta (4–8 Hz), alpha (8–12 Hz), and beta (12–30 Hz) frequency bands in the frontal, central– centroparietal, and posterior regions of interest at baseline (t0) and post-intervention or follow-up (t1). Light and dark blue bars represent the active stimulation group (n = 5), and light and dark orange bars represent the control group (n = 5), at t0 and t1, respectively. Bars indicate group means, and error bars represent the standard error of the mean (SEM) on the natural-log-transformed scale.

## Discussion

The main exploratory finding was a group-specific increase in resting-state theta power after sequential rTMS and tACS. The active group showed increased theta from baseline to post-intervention in frontal, central-centroparietal, and posterior regions, while the control group showed no comparable change. Although the Time × Group interaction was significant in the combined model and in all three separate regional analyses, the Time × ROI × Group interaction was not significant, providing no evidence of a preferential regional response. A significant Time × Group interaction was also observed for delta power, with a reduction in the control group but no significant change in the active group. No significant Time × Group interactions emerged for alpha or beta. Resting-state EEG findings in fibromyalgia are heterogeneous, although a recent review suggested an overall tendency toward reduced low-frequency activity, including theta (Silva-Passadouro et al., 2024). The present findings identify theta as a potential neurophysiological correlate of the combined intervention, but they should not be interpreted as normalization or clinical benefit.

The functional meaning of the theta increase is unclear. Higher theta has been linked to greater tenderness, fatigue, and disease impact in some studies (Diniz et al., 2024; Fallon et al., 2018), although the relationship between resting-state theta activity and clinical characteristics has varied across studies (González-Roldán et al., 2016; Lim et al., 2016; Vanneste et al., 2017). This suggests that its significance depends on region, measure, and outcome rather than reflecting a uniformly adaptive or maladaptive process. Differences in methodology, such as relative versus absolute power, further limit comparison, and cross-sectional group differences do not necessarily inform the meaning of longitudinal intervention-related change. Because we did not examine correlations between EEG changes and clinical outcomes, the increase cannot be considered a biomarker of analgesia or symptom improvement.

The theta increase was observed across all three scalp ROIs, extending beyond the sensorimotor electrodes. Previous neuroimaging studies have identified altered functional connectivity involving pain-related and sensorimotor networks in fibromyalgia (Cifre et al., 2012; Flodin et al., 2014), while changes in pain-related functional connectivity following motor-cortex stimulation have also been reported with tDCS and rTMS (Cummiford et al., 2016; Argaman et al., 2022a). However, scalp topographies do not uniquely identify neural generators (Michel & Brunet, 2019), and the present results do not establish whether the observed theta changes reflect modulation of specific cortical regions or distributed networks. The effect also emerged in the theta band despite 10-Hz stimulation. Post-stimulation effects are not necessarily confined to the stimulation frequency and may reflect plasticity-related processes rather than simple entrainment (Helfrich et al., 2014; Vossen et al., 2015), and effects outside the applied frequency have been reported previously (Bernardi et al., 2021; Tzvi et al., 2022). The null alpha finding may reflect a mismatch between the fixed 10-Hz frequency and individual mu rhythms, which we did not measure. It is also in line with a previous report of no aftereffects of 10-Hz tACS on sensorimotor alpha oscillations (Lafleur et al., 2021).

Several mechanistic limitations apply. Responses to stimulation depend on prior activity and stimulation history (Ridding & Ziemann, 2010), but the present design lacked rTMS-only and tACS-only conditions and an intermediate EEG assessment. We therefore cannot determine whether rTMS influenced the subsequent tACS response, and the theta increase should be considered in relation to the complete sequential intervention rather than as evidence of metaplasticity, additivity, or synergy. Likewise, because significant effects in one band and nonsignificant effects in others do not demonstrate frequency specificity unless compared directly (Gelman & Stern, 2006; Nieuwenhuis et al., 2011), the findings identify a prominent theta-band change but do not establish selective theta modulation, particularly given the additional Time × Group interaction observed for delta power. The physiological significance of the delta reduction in the control group is also uncertain, and the absence of a significant reduction in the active group cannot be taken as evidence that stimulation prevented such a change.

This study has further limitations. The sample was very small (n = 5 per group), so the effect sizes observed, particularly those with p values close to the significance threshold, are likely to be inflated (Gelman & Carlin, 2014); pilot data are better suited to planning future trials than to estimating effects (Bell et al., 2018; Leon et al., 2011). The no-stimulation control did not include sham stimulation and was unblinded, so nonspecific influences such as expectation, daily contact with the research team, and repeated attendance cannot be separated from stimulation effects. The ROIs were defined post hoc, and alternative electrode groupings were examined before selecting the present three-ROI configuration, increasing the uncertainty associated with regional inferences. Furthermore, rTMS exposure differed between active participants, and the groups differed at baseline in several characteristics (e.g., age and SF-36 vitality; Table 1). Four frequency bands and additional regional ANOVAs were analyzed without correction across frequency bands or separate regional analyses, increasing the risk of false-positive findings (Bender & Lange, 2001). Clinical outcomes were not reassessed after the intervention. Finally, vigilance, individual alpha peak frequency, and the aperiodic spectral component were not examined, so drowsiness, alpha slowing, or broadband changes in low-frequency power cannot be excluded as contributors to the theta increase.

In conclusion, sequential rTMS+tACS was associated with increased resting-state theta activity in fibromyalgia, but its physiological and clinical significance remains uncertain. Replication in adequately powered, sham-controlled designs with single-modality arms, intermediate EEG assessments, and EEG-clinical correlations is needed.

## Data Availability

All data produced in the present study are available upon reasonable request to the authors

## Notes

### Competing Interest Statement

The authors have declared no competing interest.

### Clinical Trial

NCT07888673

### Author Declarations

Ethics committee/IRB for Research with Medicinal Products of the Balearic Islands (CEIm-IB) gave ethical approval for this work (IB 6110/26-PI)

### Summary of Updates

The new version has been updated to include a reanalysis of the data using a frontocentral electrode pool. Although the overall findings remain similar, this revised approach incorporates a larger number of electrodes. The original figure has also been improved, and a new figure illustrating the topographical distribution of the observed effects has been added.

### Funding

- MICIU/AEI/10.13039/501100011033 and FEDER-EU: PID2022- 383 140561NB-I00

