## Supplemental Table 1 for "Sequential rTMS-tACS Neuromodulation Is Associated with Increased Resting-State Theta Activity in Fibromyalgia: A Randomized Controlled Pilot Study"

**Supplementary Material**

**Table 1**

*Sociodemographic and clinical characteristics of participants with fibromyalgia*

| **Characteristic** | **Total sample (N = 10)** | **Active stimulation (n = 5)** | **Control (n = 5)** |
| --- | --- | --- | --- |
| **Sociodemographic characteristics** |  |  |  |
| Age, years | 50.5 ± 9.1 | 46.4 ± 11.6 | 54.6 ± 3.0 |
| Age range, years | 28–58 | 28–58 | 50–58 |
| Female, n (%) | 9 (90.0) | 4 (80.0) | 5 (100.0) |
| Male, n (%) | 1 (10.0) | 1 (20.0) | 0 (0.0) |
| **Clinical characteristics** |  |  |  |
| FIQ total score | 74.38 ± 14.43 | 78.61 ± 8.41 | 70.15 ± 18.78 |
| BDI-II total score | 20.00 ± 13.23 | 17.00 ± 8.60 | 23.00 ± 17.25 |
| STAI state anxiety score | 29.40 ± 12.46 | 31.00 ± 10.86 | 27.80 ± 14.99 |
| STAI trait anxiety score | 31.40 ± 12.82 | 35.40 ± 10.90 | 27.40 ± 14.52 |
| **Alexithymia (TAS-20)** |  |  |  |
| Total score | 51.80 ± 17.27 | 55.00 ± 15.81 | 48.60 ± 19.88 |
| Difficulty identifying feelings (DIF) | 18.10 ± 8.69 | 19.60 ± 8.26 | 16.60 ± 9.79 |
| Difficulty describing feelings (DDF) | 13.70 ± 5.46 | 13.80 ± 3.96 | 13.60 ± 7.16 |
| Externally oriented thinking (EOT) | 20.00 ± 4.81 | 21.60 ± 4.72 | 18.40 ± 4.83 |
| **Pain (BPI)** |  |  |  |
| Pain severity | 7.05 ± 1.23 | 7.15 ± 0.49 | 6.95 ± 1.78 |
| Pain interference | 7.57 ± 1.12 | 7.63 ± 0.76 | 7.51 ± 1.49 |
| **Health-related quality of life** |  |  |  |
| Physical functioning | 35.00 ± 23.57 | 33.00 ± 11.51 | 37.00 ± 33.28 |
| Role limitations due to physical health | 17.50 ± 31.29 | 5.00 ± 11.18 | 30.00 ± 41.08 |
| Bodily pain | 19.40 ± 16.68 | 13.20 ± 9.23 | 25.60 ± 21.09 |
| General health | 26.70 ± 10.75 | 22.00 ± 10.37 | 31.40 ± 9.86 |
| Vitality | 31.50 ± 23.81 | 13.00 ± 5.70 | 50.00 ± 19.69 |
| Social functioning | 45.00 ± 24.44 | 35.00 ± 20.54 | 55.00 ± 25.92 |
| Role limitations due to emotional problems | 40.00 ± 30.63 | 40.00 ± 36.52 | 40.00 ± 27.89 |
| Mental health | 52.40 ± 26.56 | 43.20 ± 27.91 | 61.60 ± 24.43 |

**Table 1 (continued)**

| **Characteristic** | **Total sample (N = 10)** | **Active stimulation (n = 5)** | **Control (n = 5)** |
| --- | --- | --- | --- |
| **WHYMPI subscale scores** |  |  |  |
| Perceived social support | 4.73 ± 1.18 | 4.87 ± 1.39 | 4.60 ± 1.09 |
| General distress in the previous week | 4.03 ± 1.04 | 4.25 ± 0.77 | 3.80 ± 1.30 |
| Changes in personal relationships | 4.03 ± 1.93 | 5.13 ± 0.50 | 2.93 ± 2.25 |
| Pain interference with activity | 5.18 ± 0.60 | 5.15 ± 0.63 | 5.20 ± 0.65 |
| Changes in social activities | 4.83 ± 0.98 | 5.00 ± 0.64 | 4.65 ± 1.29 |
| Perceived self-control | 4.00 ± 0.88 | 3.40 ± 0.65 | 4.60 ± 0.65 |
| Distracting responses | 4.60 ± 1.02 | 4.30 ± 1.14 | 4.90 ± 0.90 |
| Helping responses | 3.56 ± 1.22 | 3.20 ± 1.11 | 3.92 ± 1.33 |
| Negative responses | 1.50 ± 1.18 | 1.47 ± 1.02 | 1.53 ± 1.45 |
| Household tasks | 2.57 ± 1.59 | 2.33 ± 1.31 | 2.80 ± 1.96 |
| Social activities | 2.17 ± 1.23 | 1.53 ± 0.90 | 2.80 ± 1.26 |
| Activities outside the home | 2.46 ± 0.97 | 1.68 ± 0.23 | 3.24 ± 0.74 |
| Home maintenance tasks | 1.40 ± 1.34 | 1.00 ± 1.18 | 1.80 ± 1.50 |

*Note.* Values are presented as mean ± SD, except sex, which is reported as n (%), and age ranges. Percentages use the sample size shown in each column as the denominator. All continuous measures have complete data (N = 10; n = 5 per group).

Higher scores indicate greater fibromyalgia impact (FIQ), depressive symptoms (BDI-II), anxiety (STAI), alexithymia (TAS-20), and pain severity or interference (BPI). Higher health-related quality-of-life domain scores indicate better health. The 13 WHYMPI dimensions follow the Spanish adaptation; higher scores reflect more of each named construct. Its changes in relationships and social activities refer to the effects of pain, not changes between study assessments.

BDI-II = Beck Depression Inventory-II; BPI = Brief Pain Inventory; FIQ = Fibromyalgia Impact Questionnaire; SD = standard deviation; STAI = State-Trait Anxiety Inventory; TAS-20 = 20-item Toronto Alexithymia Scale; WHYMPI = West Haven–Yale Multidimensional Pain Inventory.
